# Design and Performance Characteristics of the Elecsys Anti-SARS-CoV-2 S assay

**DOI:** 10.1101/2022.07.04.22277103

**Authors:** Karin Taffertshofer, Mirko Walter, Peter Mackeben, Julia Kraemer, Sergej Potapov, Simon Jochum

**Affiliations:** Research and Development Immunoassays; Roche Diagnostics GmbH, Penzberg, Germany; Biostatistics & Data Science, Roche Diagnostics GmbH, Penzberg, Germany

**Author notes:** **Correspondence:** Simon Jochum, Roche Diagnostics GmbH, Nonnenwald 2, 82377 Penzberg, Germany. at the time of analysis (Julia Kraemer).

**Keywords:** SARS-CoV-2, COVID-19, quantitative serology, neutralization, sensitivity, specificity

## Abstract

**Background:** Automated, high throughput assays are required to quantify the immune response after infection with or vaccination against severe acute respiratory syndrome coronavirus 2 (SARS-CoV-2). This study on the Roche Elecsys^®^ Anti-SARS-CoV-2 S (ACOV2S) assay provides insights on the assay design and performance.

**Methods:** The ACOV2S assay quantifies antibodies to the receptor-binding domain of the SARS-CoV-2 spike protein. The assigned units and the underlying standardization were compared to the international reference standard in BAU/mL. Assay specificity was assessed in samples (n=5981) collected prior to the COVID-19 pandemic and in samples from patients with non-COVID-19 respiratory infections (n=697) or other infectious diseases (n=771). Sensitivity was measured in 1313 samples from patients with mild COVID-19 and 297 samples from patients hospitalized with COVID-19. Comparison of results was performed to a comparator semi-quantitative anti-S1 assay of indirect detection format as well as a commercially available and an in-house version of a surrogate neutralization assay (ACE2-RBD).

**Results:** The originally assigned units for the ACOV2S assay were shown to be congruent to the units of the First International WHO Standard for anti-SARS-CoV-2 immunoglobulins. Overall specificity was 99.98% with no geographical differences noted and no loss of specificity in samples containing potentially cross-reacting antibodies. High sensitivity was observed, with 98.8% of samples reported to be reactive >14 days after infection and sustained detection of antibodies over time. For all samples, ACOV2S titers and neutralization capacities developed with comparable dynamics. Robust standardization and assay setup enable excellent reproducibility of results, independent of lot or analyzer used.

**Conclusion:** The results from this study confirmed that ACOV2S is a highly sensitive and specific assay and correlates well with surrogate neutralization assays. The units established for ACOV2S are also interchangeable with the units of the First International WHO Standard for anti-SARS-CoV-2 immunoglobulins. Worldwide availability of the assay and analyzers render ACOV2S a highly practical tool for population-wide assessment and monitoring of the humoral response to SARS-CoV-2 infection or vaccination.

## 1. Introduction

The assessment of the humoral immune response to infection with respiratory viruses that cause common cold-type diseases is not necessarily a typical diagnostic request. However, with the emergence in late 2019 of SARS-CoV-2, a virus that can cause severe acute respiratory syndromes, a need for serologic monitoring of antibody responses arose. In the early phase of the pandemic, the highly specific and sensitive Elecsys^®^ Anti-SARS-CoV-2 N (ACOV2N; Roche Diagnostics) assay, was developed for post-acute detection of infection in order to improve our understanding of virus circulation dynamics [1].

The SARS-CoV-2 virus is associated with considerable morbidity and mortality and has placed a substantial burden on healthcare systems. As a result, the development of an effective vaccine was prioritized, as herd immunity was considered to be key for the transition from pandemic to endemic and to limit the clinical burden of COVID-19 [2]. However, prior to the emergence of SARS-CoV-2, protective immunity to infections with viruses of the *Coronaviridae* family had been considered challenging, as protection seemed to be associated with strong individual variation and, as for many respiratory diseases, appeared to be transient [2-4]. The SARS-CoV-2 spike (S) protein was considered the most promising target for protective immunity as it is the most prominent structural protein on the surface of the virus. Consequently, the majority of SARS-CoV-2 vaccine candidates focused on the S protein or derivatives thereof [5]. In addition, vaccines based on dead or attenuated viruses contained the S protein as a prominent immunogen [6].

Detection and monitoring of the humoral immune response by quantitation of antibody titers following vaccination is well established in routine diagnostic laboratories. Detected antibodies primarily reflect the degree of a mounted humoral immune response but can also be considered as a generic marker of activation of the immune system. Also the generation of immunological memory can be deduced if the test is setup to reflect antibody affinity maturation. Appropriate antigen selection and tailoring can improve correlation of the test result to the neutralization potential of the detected antibodies. Titer monitoring over time requires quantitation of the polyclonal antibody response raised by the host. This implies that the assay has to be optimized for linear dilution of a rather heterogeneous analyte. Equally important, consistency of obtained results over time requires robust assay standardization in order to ensure reproducible results independent of manufacturing lot and time.

Quantifying antibodies to the receptor-binding domain (RBD) of the SARS-CoV-2 S protein was expected to provide a good positive predictive value for neutralization. The RBD binds to the angiotensin-converting enzyme-2 (ACE2) receptor and mediates the initial step of virus fusion with the host cell [7]. Thus, antibodies against RBD have a high likelihood of interfering with binding to ACE2, i.e., they confer neutralizing effects [5]. The RBD is a well-defined subdomain of the S protein and is composed of only 222 amino acids [8]. Its size and structure rendered it a promising antigen candidate.

We developed the Elecsys Anti-SARS-CoV-2-S (ACOV2S) assay in order to quantitatively determine the humoral response to SARS-CoV-2 infection or vaccination. Here, we describe the design of the assay and present performance data, including standardization and correlation to the first international WHO standard on anti-SARS-CoV-2 immunoglobulins, specificity and sensitivity in a large cohort of samples acquired either pre-pandemic or from patients with a polymerase chain reaction (PCR)-confirmed SARS-CoV-2 infection, and correlation with a surrogate neutralization assay.

## 2. Methods

### 2.1 Study design

The performance of the Elecsys ACOV2S assay was evaluated at Roche Diagnostics (Penzberg, Germany). All samples were collected and tested in accordance with applicable regulations, including relevant European Union directives and regulations, and the principles of the Declaration of Helsinki. Anonymized frozen serum or plasma samples were used for this study and included residual samples from blood donation centers or routine laboratory diagnostics and commercially available samples. A statement was obtained from the Ethics Committee of the Landesärztekammer Bayern confirming that there are no objections to the transfer and coherent use of anonymized leftover samples. For specificity analyses, samples from blood donors (n=2713 from the USA and n=740 from Africa) or routine diagnostic samples (n=2528 from Europe) collected before December 2019, and therefore presumed to be negative for SARS-CoV-2, were tested. In addition, cross-reactivity of the ACOV2S assay was assessed in 697 samples from patients with a respiratory infection and 771 samples from patients with other infectious, auto-immune or non-infectious hepatic disease. For the sensitivity analyses, 1610 PCR-confirmed single or sequential samples from 402 different patients with a native SARS-CoV-2 infection that occurred between February and April 2020 (i.e. infection with a strain of presumed Wuhan-Hu-1 phenotype) were tested. This included 1313 samples from 159 patients with mild COVID-19 (defined as not requiring hospitalization) and 297 samples taken from 243 patients who had been hospitalized with COVID-19. All samples had a known time difference between positive PCR test and blood draw. The date of PCR diagnosis intentionally served as the reference point for infection to circumvent less reliable references to symptom onset or similar. The majority of the samples from mild cases were taken >20 days post-PCR, conversely, the majority of the samples from hospitalized patients were taken within 20 days of a positive PCR result. A representative subset of the sensitivity samples were also tested using the Siemens SARS-CoV-2 immunoglobulin G (IgG) (COV2G) semi-quantitative assay according to the manufacturer’s instructions.

### 2.2. Elecsys Anti-SARS-CoV-2 S assay

The Elecsys Anti-SARS-CoV-2 S (ACOV2S) immunoassay is a quantitative electrochemiluminescence immunoassay (ECLIA) that detects antibodies to the SARS-CoV-2 S protein RBD. The assay applies the double antigen sandwich format detecting immunoglobulins from the sample when bridged between two specifier antigens provided by the assay. Results are automatically read-off a lot specific standard curve and reported as the analyte concentration of each sample in U/mL, with <0.80 U/mL interpreted as negative for anti-SARS-CoV-2 S antibodies and ≥0.80 U/mL interpreted as positive for anti-SARS-CoV-2 S antibodies. Values between 0.40–250 represent the linear range. Samples above 250 U/mL can be automatically diluted into the linear range of the assay with Diluent Universal (Roche Diagnostics, Rotkreuz, Switzerland). The analyzer then multiplies diluted results with the dilution factor. Experimental setups as well as data acquisition and evaluation followed standard operation procedures, which were in alignment with the respective CLSI guidelines.

### 2.3 Comparator assays

Samples were also analyzed using a comparator immunoassay and a neutralizing assay. The Siemens SARS-CoV-2 IgG (COV2G) antibody test is an automated two-step indirect chemiluminescent sandwich immunoassay that detects IgG antibodies against the spike RBD of the SARS-CoV-2 virus. Here, the detected antibodies bind with one or both of its paratopes to the immobilized antigen, while counterstaining with anti-human IgG then indirectly detects the bound antibodies. Indirect detection methods are less capable to reflect antibody maturation compared to double antigen sandwich formats [9].

COV2G results are reported in index values, with <1.0 interpreted as non-reactive (negative) for anti-SARS-CoV-2 antibodies and ≥1.0 interpreted as reactive (positive) for anti-SARS-CoV-2 antibodies [10]. Of note, the first generation COV2G assay was used in this study. This has since been replaced with an updated version with broadened dynamic range, but still utilizes an indirect detection format.

The GenScript cPass™ SARS-CoV-2 neutralization antibody detection kit is a blocking enzyme-linked immunosorbent assay (ELISA) intended for the qualitative and semi-quantitative direct detection of immunoglobulins that neutralize the interaction between RBD and hACE2. In samples that do not contain SARS-CoV-2 neutralizing antibodies, RBD conjugated to HRP can bind to ACE-2 without any impairment generating a strong signal comparable to the reference reaction. If SARS-CoV-2 neutralizing antibodies are present, they will bind to RBD-HRP and prevent the interaction with ACE-2, resulting in impaired signal. Results are reported as ratio of sample result to reference result which equals percentage binding inhibition, with <30% interpreted as negative and ≥30% interpreted as positive for SARS-CoV-2 neutralizing antibodies [11].

We also developed an in-house version of a surrogate neutralization assay for application on cobas e analyzers (Elecsys ACE2-RBD assay). Similar to cPass, but performed in solution rather than solid phase, this method measures the potential of samples containing SARS-CoV-2 antibodies to compete with ACE2-RBD binding.

#### Interference Testing

Potential interference was investigated for commonly used drugs [12, 13], drugs often used in clinical treatment of acute COVID-19 and drugs that may interrupt the RBD-ACE2 interface [14]. Potentially unspecific interference and dilution effects of the diluent used for solubilization of the drug were compensated by addition of the respective diluent without drug to a reference sample.

### 2.4 Statistical analyses

The software R, version 3.4.0 was used for comparisons to the First International WHO Standard for anti-SARS-CoV-2 immunoglobulins and for the assessment of reproducibility of results across different aliquots, days, analyzers and assay lots. All other analyses were performed using GraphPad Prism 9. For sensitivity, specificity, and precision, point estimates and 95% confidence intervals were calculated. RBD titers from sequential samples were displayed as spaghetti and smoothened median curves for the ACOV2S and comparator assays. Relative recovery was calculated for each participant as the titer measured at the last timepoint as a percentage of the highest titer measured. Qualitative agreement between the Genscript cPass and Elecsys ACE2-RBS neutralization assays was analyzed by positive predictive value (PPV) and relative specificity and sensitivity with exact 95% binomial CIs.

## 3. Results

### 3.1 Selection of antigen and assay format

The RBD antigen could be expressed in high yields and reproducibly high quality and was highly amenable to purification and labeling procedures. We also evaluated trimeric spike and monomeric spike S1 subdomain antigens, but these could be expressed with lower yields, only, and additionally required higher purification stringency. Moreover, an initial functional assessment with pre-pandemic samples suggested that better specificity was achieved for antibodies targeting the RBD compared with the trimeric spike or monomeric spike S1 antigens (**Figure 1**).

**Figure 1.**
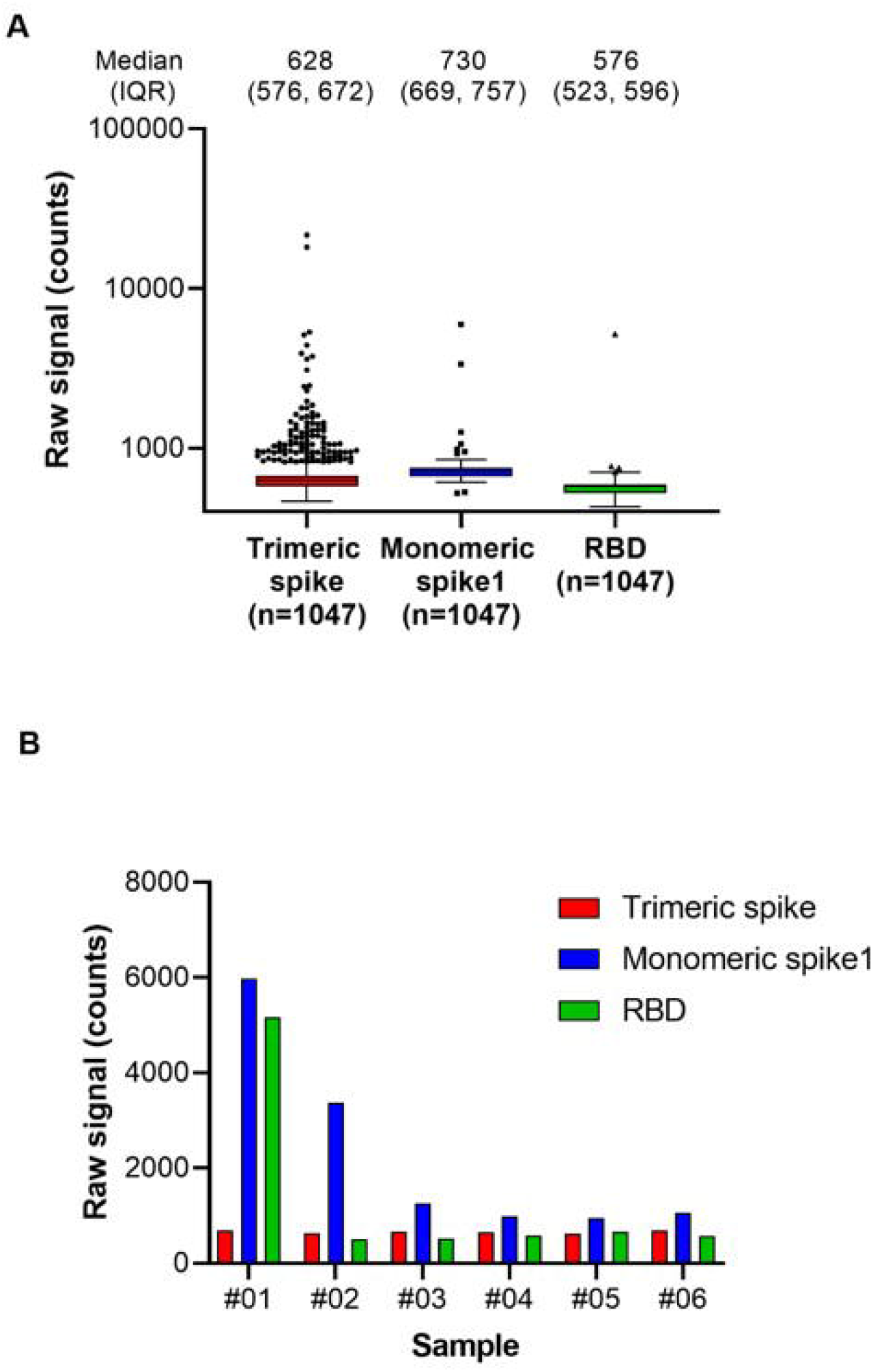
Antigen specificity in pre-pandemic serum samples (n=1047).

The Double Antigen Sandwich (DAGS) immunoassay format is well established for the detection of immunoglobulins using Elecsys assays on cobas e analyzers. Elecsys assays have a short turnaround time of typically 18 minutes, which requires a highly sensitive detection system and, most importantly, rapid formation of detectable immunocomplexes including the analyte. Successful complex formation requires the provision of all assay components in solution (no precoated solid phase) to benefit from free diffusion. This supports low thresholds for detectable concentrations and, together with the sensitive electrochemiluminescence (ECL) detection method coming with a broad linear range, enables strong signal dynamics of the DAGS method used in Elecsys assays. The DAGS format per se does not differentiate between immunoglobulin classes. However, bridging of two antigens is required for signal generation, which leads by design to the predominant detection of high-affinity antibodies, i.e. IgG. We sought to further sustain this tendency by the provision of well-defined and monomeric antigens as well as rather stringent buffer conditions, such that single binding events, as mediated by IgG, are the major signal driver. RBD was ultimately selected as the ideal candidate to fulfill these requirements. By design, DAGS assays feature strong specificity and do not require sample predilution to reduce unspecific reactivity. The application of a highly specific antigen further corroborates this feature. Together with the adjustment of buffer stringency, these aspects serve to balance the required specificity versus sensitivity.

### 3.2 Standardization and correlation to WHO standard

At the time of the development and launch of the ACOV2S assay, no standard reference material existed for qualitative SARS-CoV-2 antibody assays. We developed an internal reference standard to enable reproducible and lot independent test results in arbitrary units. Parallel assessment of a medical decision point (MDP) for differentiation of samples being non-reactive or reactive for RBD-specific antibodies indicated that such an MDP might lie close to the numeric unit of 1 on this scale. In addition, established working calibrators based on pools of native human sample material were adjusted to cover critical supporting points enabling the generation of a robust standard curve. The primary measuring range of the ACOV2S assay was defined from 0.4 to 250 U/mL, determined by optimal linear detection of a dilution series of human samples and with the majority of result within this range following native infection with a mild course of disease. While retrospectively a larger measuring range had been desirable, automated dilution provides a convenient workaround. We subsequently observed that the units of the First International WHO Standard for anti-SARS-CoV-2 immunoglobulins (NIBSC code 20/136) were congruent to the originally assigned units based on the internal standard (**Figure 2**). Linearity was proven using a dilution series of the WHO standard as well as excellent value recovery. Robustness of the applied standardization procedure could be proven by the use of three different assay lots, one representing the initial assay lot undergoing reference standardization (DR lot), an additional lot from early phase of routine production (MP02) and another routine lot produced 3 production events later (MP05, 9 months after DR lot production). The data shown in figure 2 were generated by parallel analysis of these three lots. The observed congruency of the results indicates good stability of the product over time as well as reliable standardization ensuring the same numeric results independent of lot and product age.

**Figure 2.**
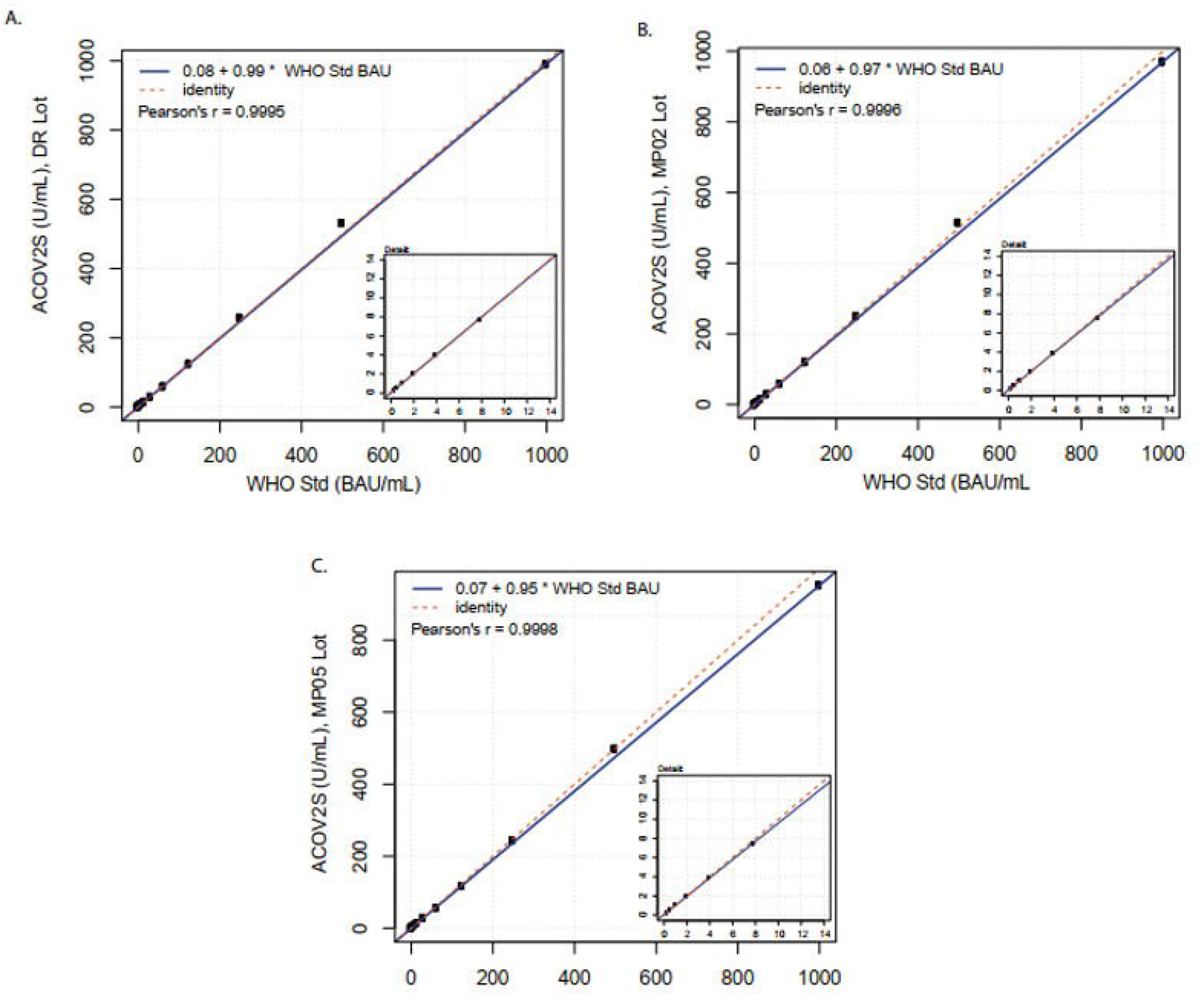
Correlation of ACOV2S units to the First International WHO Standard for anti-SARS-CoV-2 immunoglobulins. Linearity using a dilution series of the WHO standard for (A) lot DR, (B) lot MP02, and (C) lot MP05.

### 3.3 Specificity

Specificity was extensively assessed in pre-pandemic samples obtained from blood donors in the USA and Africa and routine diagnostic samples acquired in Europe. One presumably false-positive result was observed from >5000 samples in total, giving an overall specificity of 99.98%. No differences in specificity were observed between samples from different geographical locations and no differences were observed between different serological backgrounds (**Table 1**). Specificity of the ACOV2S assay was also confirmed in a cohort of pre-pandemic samples with potentially cross-reacting antibodies to related indications, i.e., respiratory diseases (**Table 2**) or less closely related clinical indications (**Table S1**). Potential assay interference from common drugs, special drugs used in COVID-19 treatment or drugs with postulated potential to interrupt the RBD-ACE2 interface [14] was also assessed. For the vast majority, no significant interference was observed as analyte recovery in spiked samples (1–3x daily dose) was within acceptance range (90–110%) compared to the unspiked reference sample (**Table S2**). Analyte recovery was 69–80% using higher drug concentrations of ritonavir (0.360 and 0.480 mg/L) and 87–88% with highest itraconazole concentration (30 mg/L).

**Table 1.** Specificity in samples obtained from routine diagnostic laboratories and blood donors pre-pandemic.

| **Sample cohort** | **Samples (n)** | **Reactive (n)** | **Specificity** | **LCL (95%)** | **UCL (95%)** |
| --- | --- | --- | --- | --- | --- |
| Diagnostic routine EU | 2528 | 0 | 100.00 % | 99.85 % | 100.00 % |
| Blood donations USA | 2713 | 1 | 99.96 % | 99.79 % | 100.00 % |
| Blood donations Africa | 750 | 0 | 100.00 % | 99.51 % | 100.00 % |
| **Overall** | **5991** | **1** | **99.98 %** | **99.91 %** | **100.00 %** |

LCL, lower confidence limit; UCL, upper confidence limit.

**Table 2.**
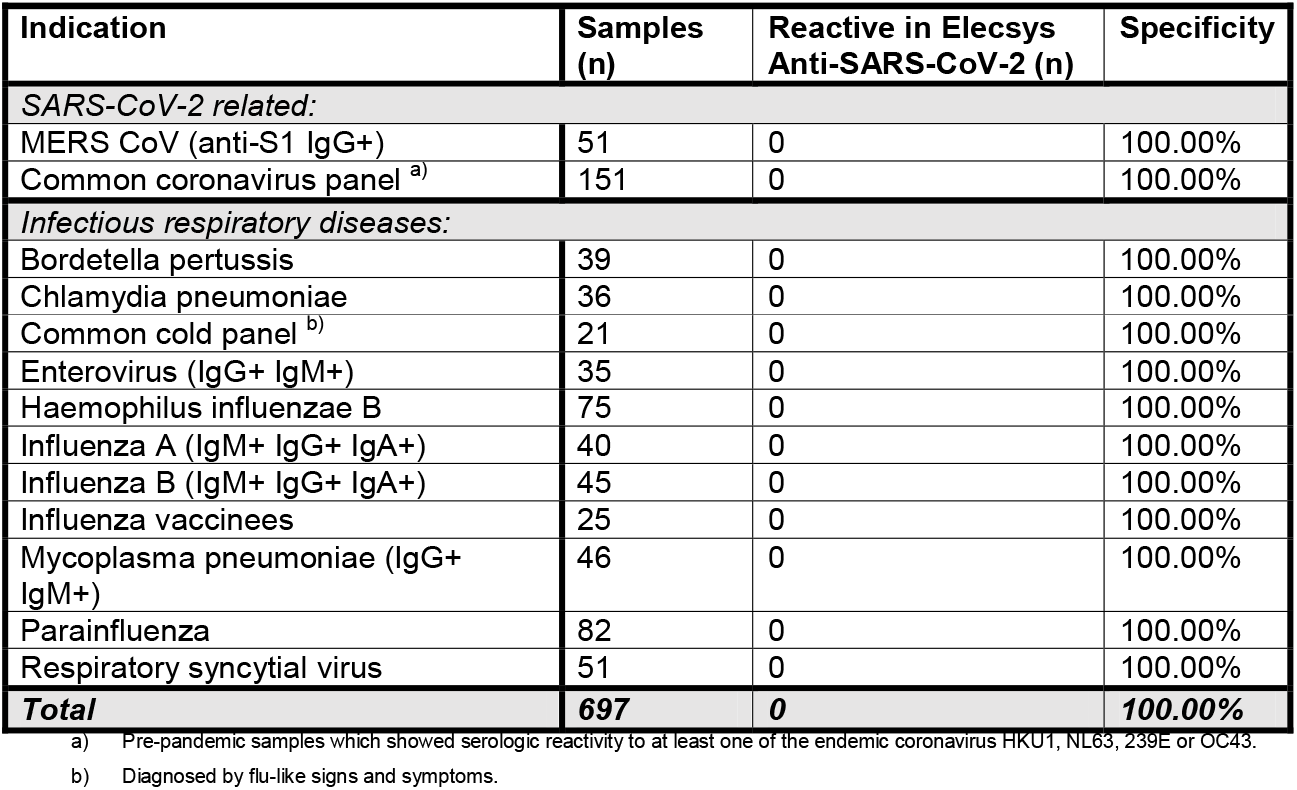
Cross-reactivity of the ACOV2S assay in samples from participants with respiratory infections.

### 3.4 Sensitivity

Sensitive detection of seroconversion was observed for patients with a native infection with SARS-CoV-2, with most samples determined “reactive” by ACOV2S within 14 days of a positive PCR test. Sensitivity increased with the onset of the humoral immune response, with 98.8% of samples reported to be reactive >14 days after infection. By day 28, all samples for patients in this cohort were determined to be reactive (**Figure 3**).

**Figure 3.**
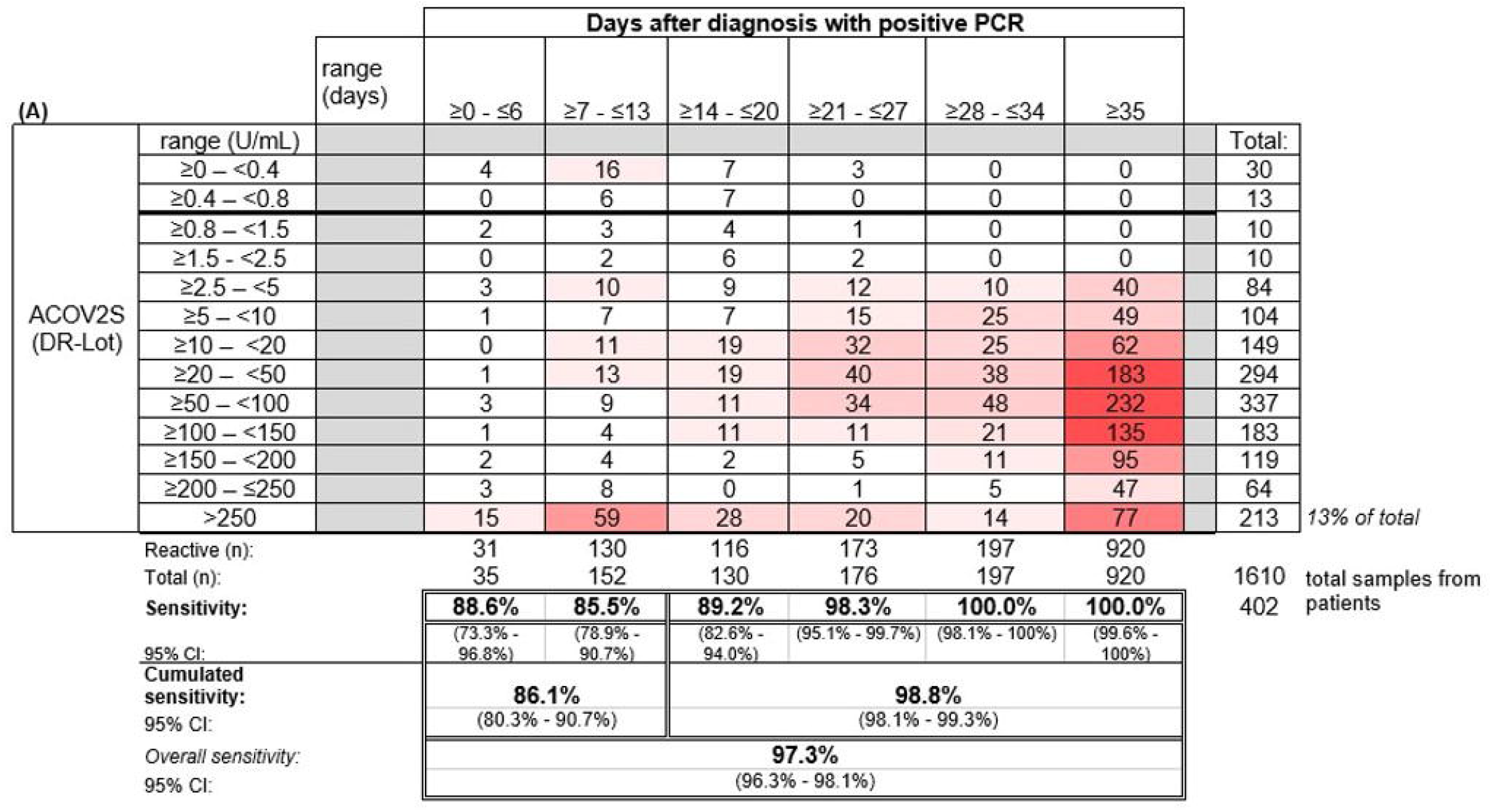
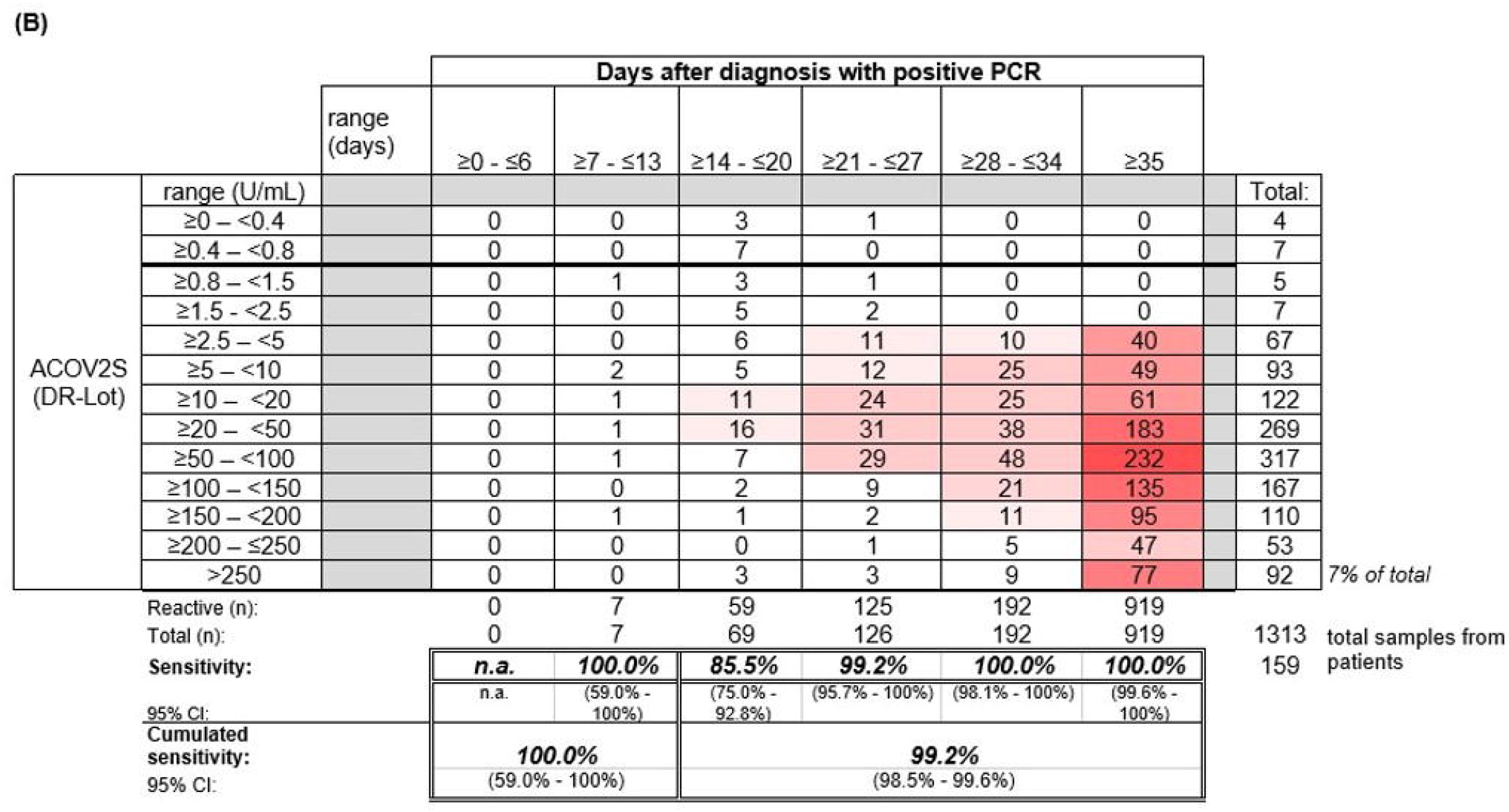
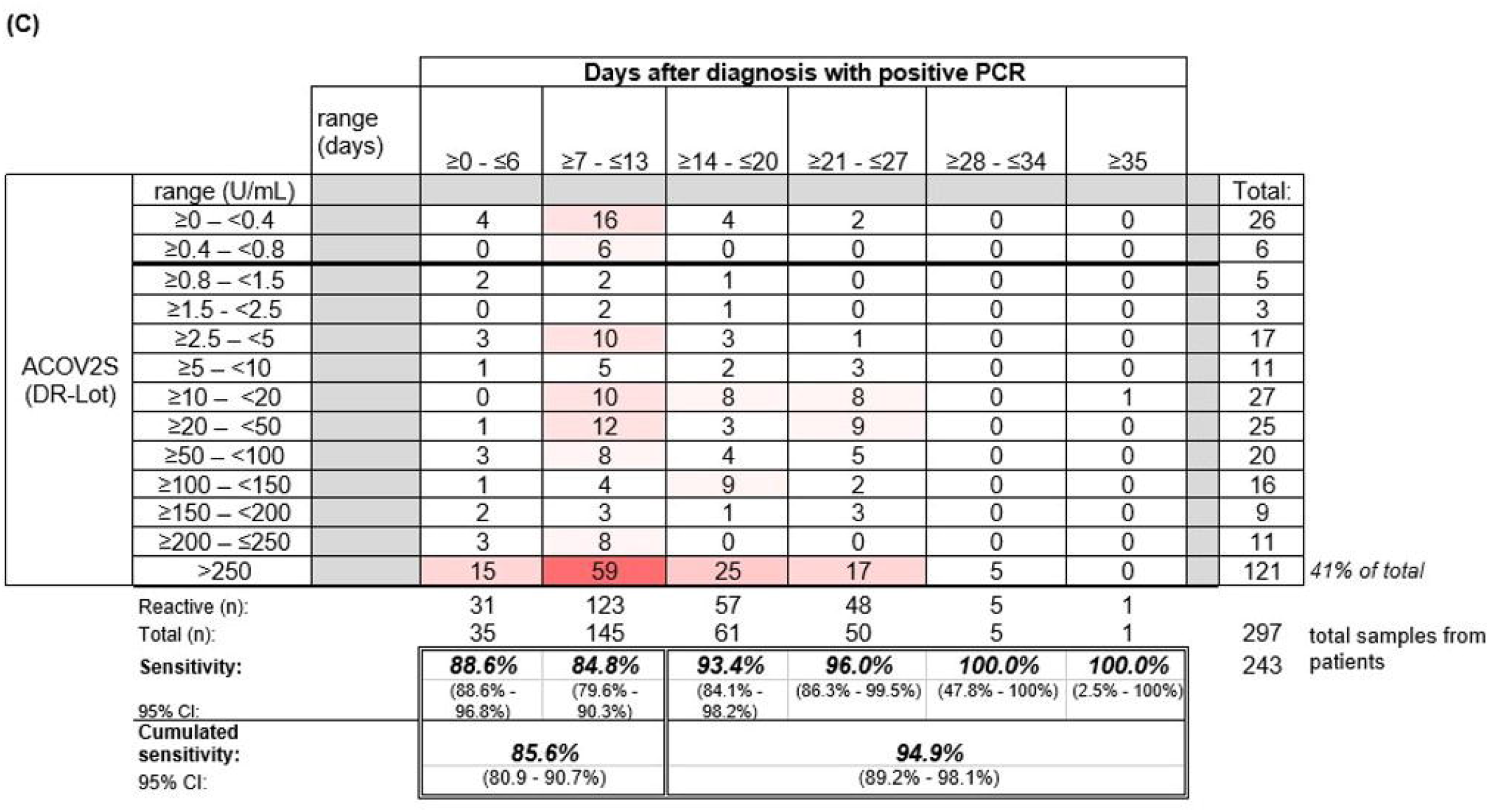
Sensitivity of the ACOV2S assay. (A) Sensitivity of the ACOV2S assay in samples from (A) all patients, (B) patients with mild symptoms of COVID-19, and (C) patients hospitalized due to COVID-1.

Samples from patients with mild COVID-19 exhibited average rising titers over time as indicated by the heat map (**Figure 3B**). In total, only 7% of all samples from patients with mild disease exceeded the primary measuring range (>250 U/mL). The patient with the latest seroconversion was part of the mild disease cohort. For patients with severe disease, i.e., hospitalized patients, higher titers were observed early after diagnostic PCR, with 41% of all samples exceeding the primary measuring range (most of them exceeded 250 U/mL within 14 days after diagnostic PCR) (**Figure 3C**). Rapid development of relatively high titers was observed in case of severe course of disease, whereas moderate, still continuous antibody titer development seemed to be associated with mild disease.

A tendency for stable or even increasing anti-S titers over time following native infection was observed using the ACOV2S assay. This overall trend can be visualized in a smoothened median (50-day average) of all samples per timepoint (**Figure 4A**). Using the COV2G comparator method applying indirect antibody detection, the reverse effect was reported, i.e., antibody levels appear to be waning over time (**Figure 4A**).

**Figure 4.**
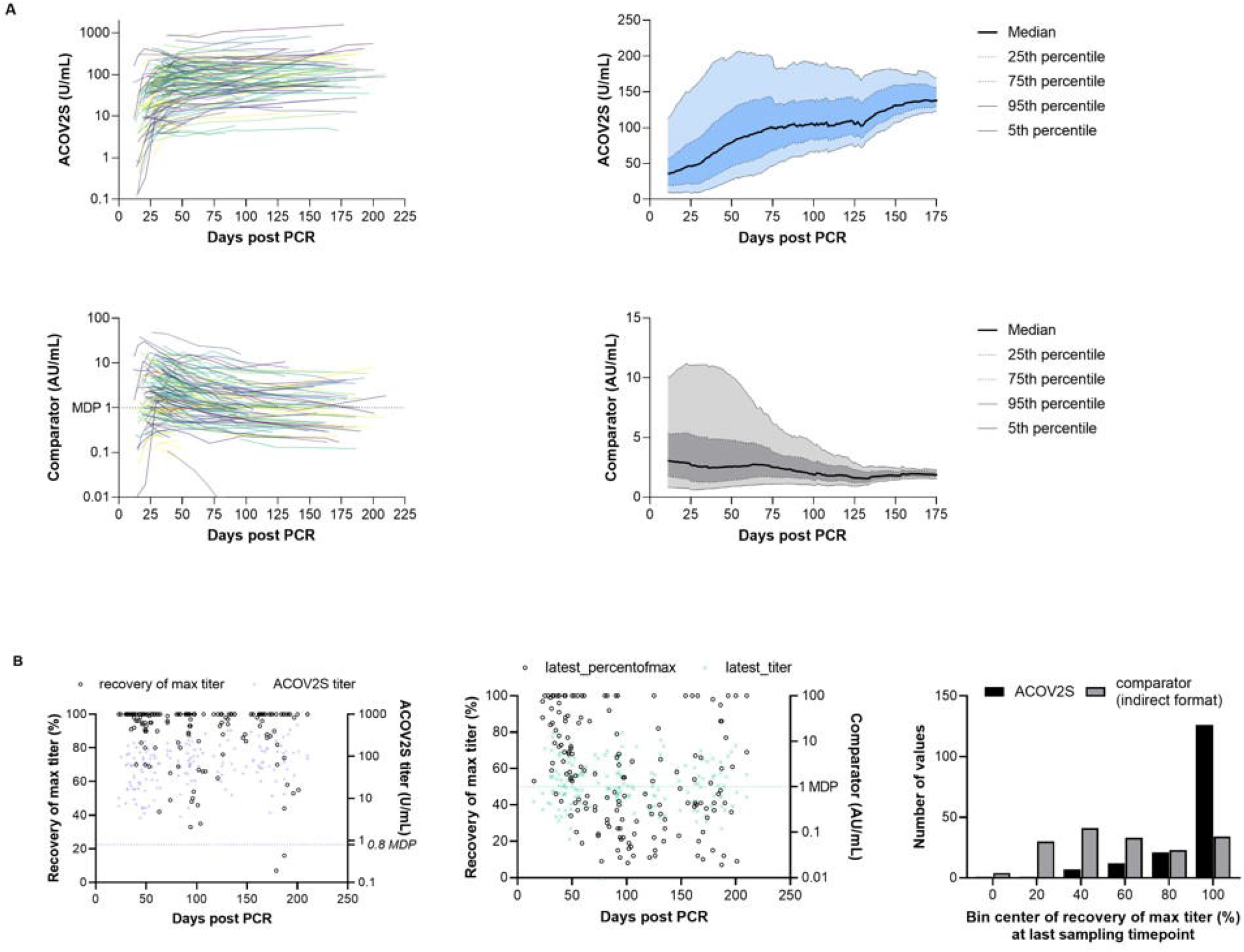
RBD titer dynamics. (A) Spaghetti and smoothened median curves for RBD titers over time for the ACOV2S and comparator assay, and (B) relative recovery at the last sampling timepoint for the ACOV2S and comparator assay (absolute titer values at the last sampling timepoint are plotted in color [right y-axis] and relative recovery is plotted in black [left y-axis].

Scales of different methods can be difficult to harmonize even when standardized to the same reference, in particular when different antigens are used for detection [15]. So absolute numeric values cannot be directly compared, but relative titer development over time are a more suitable comparator. The plots in **Figure 4B** show the titer measured at the latest sampling timepoint for an individual as a percentage of the highest value obtained for that individual (relative recovery), i.e. values with 100% recovery indicate that the highest titer value for a participant was obtained with the last sampling time point or that no titer waning from a previous timepoint had occurred. The bar chart shows the cumulative frequency distribution of results. Using the ACOV2S assay, the majority of the patients exhibited 100% relative recovery at the last sampling timepoint, indicating sustained antibody titers. In contrast, relative recovery for the comparator assay was more variable and results of this method waned in a considerable proportion of the patients.

### 3.5 Correlation with neutralization assays

The correlation of ACOV2S with a surrogate neutralization assay (cPass™, Genscript) has already been described.[16] Here, we investigated the correlation of the development of neutralizing potential over time as determined with cPass and antibody titer development as determined with ACOV2S using samples from patients with severe and mild course of disease from our sensitivity cohort. The samples included representative time points of longitudinal sample panels from individual donors with mild disease (n=22) to additionally resolve cPass and ACOV2S. In these longitudinal panels, ACOV2S titers and neutralization capacities developed with comparable dynamics (**Figure 5A and S1**). The correlation of the numeric value of inhibition as determined with cPass to the ACOV2S titer was very good in the individually resolved time buckets, despite cPass inhibition appeared less pronounced at later time points compared to ACOV2S antibody titers (**Figure 5B**). As in vitro determination of inhibition intentionally allows for saturation and hence does not reflect antibody affinity, but the ACOV2S method does, this might be a likely reason for the observed quantitative correlation dynamics. However, cPass is approved for qualitative interpretation only and the majority of ACOV2S reactive samples qualified “inhibitory” also over time. The application of 15 U/mL in ACOV2S as the decision point for correlation to inhibition led to an improved PPV of the ACOV2S result on presence of inhibition in all investigated time buckets (**Figure 5C**). In general, all samples with an ACOV2S titer of ≥150 U/mL or higher seemed to saturate the inhibition potential in cPass at all addressed time buckets (**Figure 5B**).

**Figure 5.**
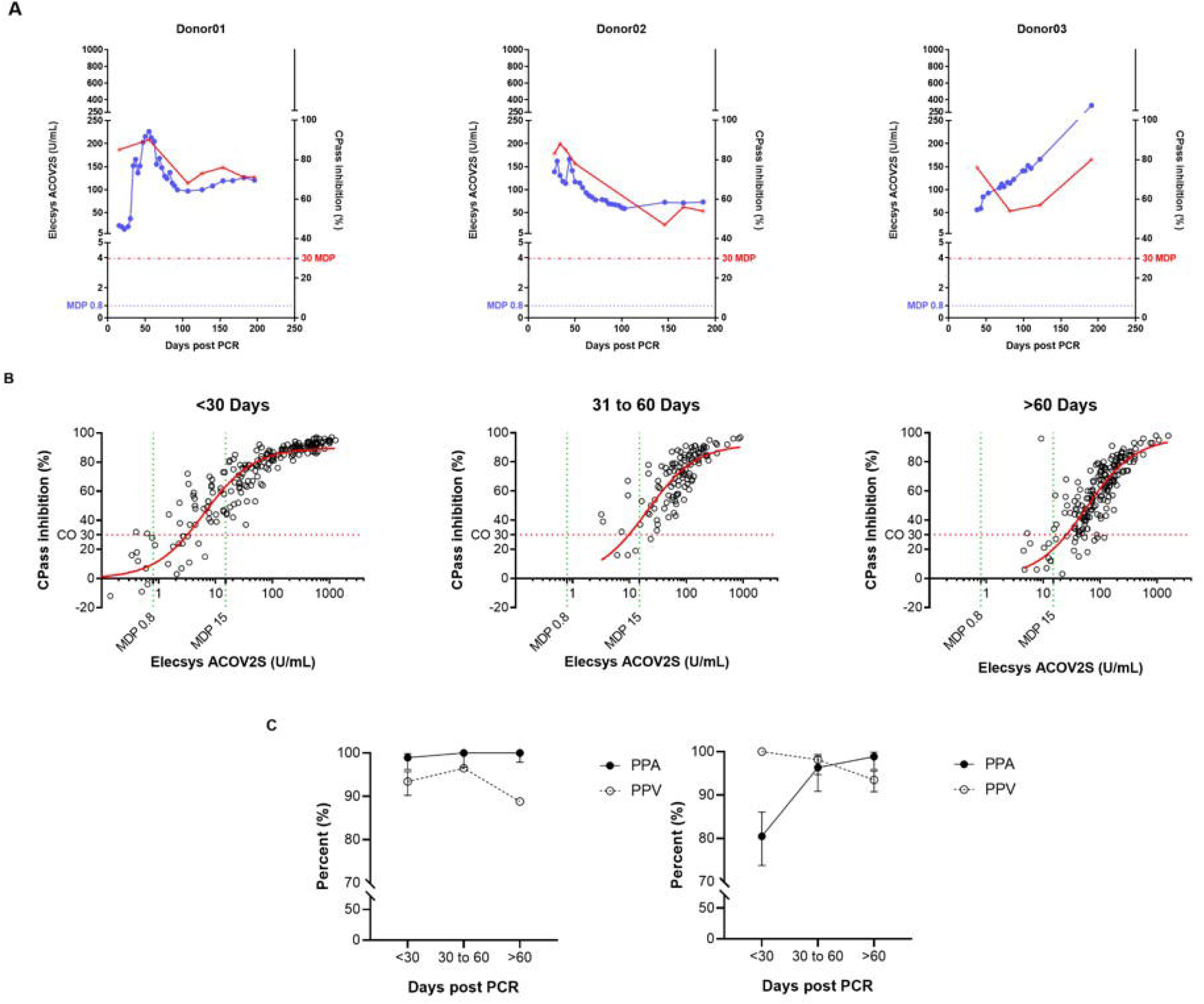
Correlation of the ACOV2S assay with results from the cPass neutralization assay. (A) ACOV2S titers and percentage inhibition from the cPass assay from longitudinal samples from exemplary selected individual donors, and (B) comparison of the ACOV2S and cPass results stratified in time buckets after diagnostic PCR and (C) derived qualitative agreements. ACOV2S reactivity in samples with cPass inhibitory capacities is shown as positive percent agreement (PPA), cPass inhibitory capacity ACOV2S reactive samples is shown as positive predictive value (PPV). Analyses were carried out applying both, 0.8 U/mL and 15 U/mL as decision point for relevant ACOV2S reactivity.

We observed a good correlation of cPass results with the in-house Elecsys ACE2-RBD surrogate neutralization assay (**Figure S2**). A qualitative cutoff of 25% inhibition in the Elecsys ACE2-RBD method indicated the presence of neutralizing activity at a comparable level to the cPass method when applying the cPass cut off of 30%. A relative sensitivity of 93.4% was determined for the Elecsys ACE2-RBD assay and a ROC analysis resulted in an area of 0.9599 under the curve (**Figure S2**).

The internal competitive assay format correlated well with the titers determined with ACOV2S (**Figure S3**). Quantitative result correlation, as well as qualitative result interpretation, showed excellent comparability of the internal ACE-RBD method with ACOV2S.

Of note, the investigated sample cohort did not include a balanced amount of defined negative samples, but was conducted with samples from patients with confirmed SARS-CoV-2 infection. For this reason, a negative predictive value was not considered as well as the accuracy of the indicated negative agreement might be limited.

### 3.6 Precision and reproducibility

The ACOV2S assay demonstrated excellent precision (both within run and within lab) independently of the analyzer used, with the high throughput analyzer of the latest generation (cobas e 801) having slightly better precision than a benchtop analyzer of the previous generation (cobas e 411) (**Table 3**). Reproducibility of the ACOV2S assay was investigated by assessment of differences in the result for aliquots of the same samples (n=5) measured on different days (n=5), on analyzers of different types (e411, e601, e801; n=3) and with different lots (n=3). In the worst case setting, only a marginal coefficient of variation was observed with 6.7% at LLOQ level and 3.2% at the high end of the primary measuring range **(Table 4)**.

**Table 3.** Precision of the ACOV2S assay using the cobas e 411 and cobas e 801 analyzers.

| Sample Material | Mean U/mL | Repeatability |  | Intermediate precision |  |
| --- | --- | --- | --- | --- | --- |
|  |  | (within-part precision) |  | (within-lab/total) |  |
|  |  | SD Estimate | CV% Estimate | SD Estimate | CV% Estimate |
| cobas e 411 |  |  |  |  |  |
| Sample1 | 0.017 | 0.009 | na | 0.009 | na |
| Sample2 (HSP1) | 0.483 | 0.014 | na | 0.016 | na |
| Sample3 (HSP2) | 0.826 | 0.023 | na | 0.023 | na |
| Sample4 (HSP3) | 5.74 | na | 2.30 | na | 2.60 |
| Sample5 (HSP4) | 12.3 | na | 2.20 | na | 2.50 |
| Sample6 (HSP5) | 54.6 | na | 2.90 | na | 2.90 |
| Sample7 (HSP6) | 77.9 | na | 2.30 | na | 2.70 |
| Sample8 (HSP7) | 190 | na | 1.60 | na | 1.90 |
| Sample9 | 260 | na | 2.30 | na | 2.50 |
| cobas e 801 |  |  |  |  |  |
| Sample1 | 0.020 | 0.015 | na | 0.015 | na |
| Sample2 (HSP1) | 0.483 | 0.014 | na | 0.014 | na |
| Sample3 (HSP2) | 0.826 | 0.015 | na | 0.015 | na |
| Sample4 (HSP3) | 5.69 | na | 2.10 | na | 2.40 |
| Sample5 (HSP4) | 12.0 | na | 1.30 | na | 1.60 |
| Sample6 (HSP5) | 54.8 | na | 1.40 | na | 1.40 |
| Sample7 (HSP6) | 77.3 | na | 1.60 | na | 2.00 |
| Sample8 (HSP7) | 184 | na | 0.900 | na | 1.40 |
| Sample9 | 253 | na | 1.30 | na | 1.70 |

**Table 4.** Reproducibility of the ACOV2S assay. For each sample, measurements were made using different aliquots, days, analyzers and lots.

|  | Sample conc. | Deter. | Repeatability |  | Between-Day |  | Between-Lot |  | Between-Device |  | Reproducibility |  |
| --- | --- | --- | --- | --- | --- | --- | --- | --- | --- | --- | --- | --- |
| Sample | Mean U/mL | N | SD | CV [%] | SD | CV [%] | SD | CV [%] | SD | CV [%] | SD | CV [%] |
| Prec02 | 0.465 | 225 | 0.0142 | n.a. | 0.0148 | n.a. | 0.0133 | n.a. | 0.0159 | n.a. | 0.0291 | n.a. |
| Prec03 | 0.47 | 225 | 0.0140 | n.a. | 0.0115 | n.a. | 0.0202 | n.a. | 0.0160 | n.a. | 0.0315 | n.a. |
| Prec04 | 0.818 | 225 | 0.0183 | n.a. | 0.0224 | n.a. | 0.0387 | n.a. | 0.0176 | n.a. | 0.0514 | n.a. |
| Prec05 | 0.94 | 225 | 0.0180 | n.a. | 0.0165 | n.a. | 0.0143 | n.a. | 0.0206 | n.a. | 0.0350 | n.a. |
| Prec06 | 5.57 | 225 | n.a. | 1.9 | n.a. | 2.0 | n.a. | 3.2 | n.a. | 1.8 | n.a. | 4.6 |
| Prec07 | 12 | 225 | n.a. | 2.1 | n.a. | 1.9 | n.a. | 4.5 | n.a. | 2.0 | n.a. | 5.7 |
| Prec08 | 53.6 | 225 | n.a. | 2.1 | n.a. | 1.6 | n.a. | 3.4 | n.a. | 1.7 | n.a. | 4.6 |
| Prec09 | 73.9 | 225 | n.a. | 2.1 | n.a. | 2.2 | n.a. | 4.5 | n.a. | 2.2 | n.a. | 5.9 |
| Prec10 | 186 | 225 | n.a. | 1.6 | n.a. | 1.7 | n.a. | 2.8 | n.a. | 2.7 | n.a. | 4.5 |
| Prec11 | 257 | 225 | n.a. | 1.8 | n.a. | 1.8 | n.a. | 0.8 | n.a. | 1.9 | n.a. | 3.3 |

These data confirm that the applied standardization ensures excellent result recovery and lot comparability as well as independence of results from the type of analyzer used.

## 4. Discussion

ACOV2S is a highly sensitive and specific assay for the detection of antibodies to the SARS-CoV-2 S protein. The RBD was chosen as the antigen for this assay as it is highly immunogenic, part of all vaccines registered so far and a prominent target for neutralizing antibodies due to its high functional relevance for the virus[5, 7]. Moreover, the use of the RBD antigen resulted in higher specificity compared to using larger antigens and the well. Also, the well-defined and relatively small RBD supports linear dilution of the polyclonal and heterogeneous antibody mixture of a sample to the best extent.

The ACOV2S assay offers both excellent sensitivity and specificity for the detection of anti-RBD antibodies following a native SARS-CoV-2 infection, as shown here, as well as following vaccination with mRNA-1273 (Spikevax; Moderna, Cambridge, MA) [17-19]. In this analysis, the high specificity of ACOV2S was confirmed in a large set of samples collected prior to the emergence of SARS-CoV-2. Strong specificity was critical in the beginning of the pandemic and low prevalence of disease and absence of vaccination. However, strong specificity is also advantageous in vaccinated populations, e.g., ruling out false-positive results and preventing non-convalescent or unvaccinated individuals from being put at risk of infection. The ACOV2S assay demonstrated no cross-reactivity to other respiratory or non-respiratory infections, including in samples from participants from Africa who are likely to have a different disease and immunological background compared to samples from Western participants. This is in contrast to reports in the literature, which identified possible false-positive SARS-CoV-2 serology (using non-ACOV2S assays) in pre-pandemic samples from malaria- or dengue fever-infected patients [20-22]. With the growing proportion of infected and, most prominently, vaccinated individuals, the strong specificity of ACOV2S is a reliable tool to identify naïve, i.e. vulnerable individuals.

Assays that measure antibodies against the RBD, such as ACOV2S, are by design expected to provide a strong positive predictive value for neutralization compared with assays that measure antibodies to the full S protein (expected to detect a large number of antibodies to other epitopes that are less likely to be neutralizing). In this and previous studies, the results from the ACOV2S assay have been shown to correlate well with *in vitro* and surrogate neutralization assays [17, 18, 23]. Application of in vitro neutralization assays with high throughput is cost and labor intense as well as laboratory sources providing this service are scarce. Also, considerable result variation is typically inevitable with assays involving life cell culture. Surrogate neutralization assays measuring interference with ACE-RBD interaction circumvent these disadvantages to a certain extent but apply a competitive assay format that typically comes with trade-offs in terms of dynamic range and robustness. The strong correlation of ACOV2S results with surrogate neutralization assays applied in this study renders the obtained ACOV2S results a suitable alternative to surrogate NT testing, with ACOV2S offering superior performance, dynamics and robustness.

No standard reference material existed for quantitative SARS-CoV-2 antibody assays at the time that the ACOV2S assay was initially developed. However, we have shown here that the units that were established for ACOV2S are interchangeable with the units of the First International WHO Standard for anti-SARS-CoV-2 immunoglobulins, signifying that data generated with ACOV2S can be assessed retrospectively without the need for restandardization.

Longitudinal monitoring of patients with mild course disease show relatively stable or rising antibody titers over time when determined with ACOV2S and in contrast to the competitor method. These findings are in line with other reports on reduced waning of antibody titers with DAGS format assays.[9, 24] Reinfection or consistent restimulation due to lack of viral clearance cannot be ruled out but would be detectable with an indirect method as well. However, DAGS format assays are able to reflect both and translate raising antibody concentration as well as raising antibody affinity into raising signals. This implies that low concentrations of high avidity antibodies can lead to the same result in the ACOV2S assay as high concentrations of low avidity antibodies; the biological function and the likelihood of detecting foreign target antigens in vivo are probably also comparable in both scenarios. In contrast, indirect formats do not differentiate if an antibody is bound with one or both paratopes to the antigen, either leads to the same signal. Antibody quality in terms of affinity is therefore more difficult reflect in an indirect format assay.

Taking the results from both formats together, the rapid decline in SARS-CoV-2 specific antibody levels as reported by indirect assays format appears to be counter-acted by rapid affinity maturation as reflected by ACOV2S and resulting stable or rising test results. Of note, the degree of antibody waning in indirect assay formats seems to exceed the loss of *in vitro* neutralization capacities and even the *in vivo* efficacy of immunity.[25-27] Although it is not strictly necessary to distinguish between immunoglobulin classes during serology screening, field observations have demonstrated that individuals may have a robust and protective immune response to re-infection that is longer lasting than indirect antibody detection formats would suggest. [28, 29] Whether the ACOV2S antibody titer shows a greater correlation with a reduced risk of reinfection remains to be investigated. It is also open as to what extent neutralization assays also reflect the important aspect of antibody affinity or if they are more reflective of antibody concentrations only. Also, the correlation of antibody mediated neutralization as measured in vitro to a reduced risk of infection or protection from severe disease is, to our knowledge, not yet reliably established.

In general, there is a risk that assay results waning below cut-off or even LOQ suggest that individuals are immune naïve and hamper titer monitoring. Differentiation of naïve patients from patients with presumed critically low immunity becomes more challenging. Following ACOV2S titer development in immunocompromised patients will enable insights on their general humoral immune response and correlation of their risk to develop severe disease and, at the same time, to get an impression about antibody maturation under immunosuppression based on the observed ACOV2S titer dynamics.

The results presented here emphasize that antibody results generated using different detection methods and/or different detection antigens should not be considered interchangeable. Despite attempts at harmonization by application of uniform reference material, the individual humoral immune response and its polyclonal nature together with individual differences in antibody maturation and selection are likely to lead to significantly different results using assays that utilize different methods of detection.

Absolute antibody titers and dynamics may be a marker of disease severity [17, 30]. In this study, in patients with mild COVID-19, antibody titers increased gradually over time. In contrast, antibody titers increased more rapidly and to a higher level in patients who were hospitalized. Although data are currently limited, some evidence is beginning to emerge supporting the medical value of antibody testing in the acute management of patients infected with SARS-SoV-2 [31].

Straightforward and well-established workflows combined with reliable detectability render antibody levels an appealing marker of immune response to viral infection or vaccination. It remains to be elucidated as to what extent they can be considered a direct protective effector or merely a surrogate marker for activation of the immune system. In both scenarios, the role of antibody dynamics should be further investigated to elaborate if certain titer limits correlate with a reduced risk of severe course of disease and which co-variates are to be taken into account, like e.g. age, immune status, and also putative additional markers. The SARS-CoV-2 pandemic, which resulted in population-wide immunization campaigns and detailed clinical characterization of the disease course, has provided a rich field to investigate possible correlations in detail and for robust statistics to be generated. Emerging new variants of the SARS-CoV-2 virus add an additional level of complexity to these attempts. However, obtaining reliable numeric results over time is key to additionally taking this layer into account by putative correction factors, adaptation of thresholds and adjusting the expectable level of confidence of predictive values (although currently still theoretical). In this study, the ACOV2S assay demonstrated excellent precision and reproducibility, which are prerequisites for reliable titer monitoring over time. Virtually worldwide availability of the assay and analyzers render ACOV2S a suitable candidate for population-wide assessment and monitoring of the humoral response elicited by the SARS-CoV-2 S virus.

## 5. Conclusion

ACOV2S is a highly sensitive and specific assay for the detection of antibodies to the RBD of the SARS-CoV-2 S protein. In this analysis, we have demonstrated standardization and correlation to the first international WHO standard on anti-SARS-CoV-2 immunoglobulins and correlation with surrogate neutralization assays. An analysis of absolute antibody titers and dynamics suggested that the assay can have value in determining disease severity. Furthermore, the DAGS format allows for the detection of low concentrations of high avidity antibodies, meaning that the ACOV2S assay may retain sensitivity during the process of antibody maturation. The ability of ACOV2S to predict the risk of severe disease in vaccinated or convalescent patients remains unknown but is of great interest in the current phase of the pandemic.

## Data Availability

The study was conducted in accordance with applicable regulations. For more information on the study and data sharing, qualified researchers may contact the corresponding author.

## Acknowledgements

ELECSYS and COBAS are trademarks of Roche. All other product names and trademarks are the property of their respective owners. The Elecsys Anti-SARS-CoV-2 S assay is approved under Emergency Use Authorisation in the US. Editorial support was provided by Steph Carter and Jade Drummond of inScience Communications, Springer Healthcare Ltd, UK, and was funded by Roche Diagnostics International Ltd (Rotkreuz, Switzerland).

## Author contributions

Study concept/design: SJ, MW, PM, KT

Data acquisition: KT, JK, MW, PM

Data analysis and interpretation: KT, MW, PM, JK, SJ, SP

Review and final approval of manuscript: SJ, MW, KT, SP

