## Supplementary material for "Design and Performance Characteristics of the Elecsys Anti-SARS-CoV-2 S assay"

**Supplementary Table S1. Cross-reactivity of the ACOV2S assay in samples from patients with non-respiratory infections.**

| **Indication** | | | **Samples (n)** | **Reactive in Elecsys Anti-SARS-CoV-2 (n)** | **Specificity** |
| --- | --- | --- | --- | --- | --- |
| ***Other infectious diseases:*** | | | | | |
| Adenovirus | | | 25 | 0 | 100.0% |
| Borrellia | | | 6 | 0 | 100.0% |
| Candida albicans | | | 13 | 0 | 100.0% |
| Chlamydia trachomatis | | | 12 | 0 | 100.0% |
| CMV acute (IgM+ IgG+) | | | 86 | 0 | 100.0% |
| E.coli (anti-E.coli reactive) | | | 10 | 0 | 100.0% |
| EBV acute (IgM+ VCA IgG+) | | | 106 | 0 | 100.0% |
| Gonorrhea (Tripper) | | | 5 | 0 | 100.0% |
| HAV acute (IgM+) | | | 10 | 0 | 100.0% |
| HAV late (IgG+) | | | 15 | 0 | 100.0% |
| HAV vaccinees | | | 15 | 0 | 100.0% |
| HBV acute | | | 12 | 0 | 100.0% |
| HBV chronic | | | 12 | 0 | 100.0% |
| HBV vaccinees | | | 15 | 0 | 100.0% |
| HCV | | | 50 | 0 | 100.0% |
| HEV | | | 12 | 0 | 100.0% |
| HIV infection | | | 10 | 0 | 100.0% |
| HSV acute (IgM+) | | | 24 | 0 | 100.0% |
| HTLV | | | 6 | 0 | 100.0% |
| Legionella (IgGAM+) | | | 7 | 0 | 100.0% |
| Listeria | | | 6 | 0 | 100.0% |
| Measles | | | 10 | 0 | 100.0% |
| Mumps | | | 14 | 0 | 100.0% |
| Parvovirus B19 | | | 30 | 0 | 100.0% |
| Plasmodium falciparum (Malaria) | | | 8 | 0 | 100.0% |
| Rubella acute (IgM+, IgG+) | | | 12 | 0 | 100.0% |
| Toxoplasma gondii (IgM+, IgG+) | | | 8 | 0 | 100.0% |
| Treponema pallidum (Syphilis) | | | 62 | 0 | 100.0% |
| VZV (Varicella zoster) | | | 30 | 0 | 100.0% |
| ***Auto-immune diseases:*** | | | | | |
| AMA (anti-mitochondrial antibodies) | | | 30 | 0 | 100.0% |
| ANA (anti-nuclear antibodies) | | | 17 | 0 | 100.0% |
| Hemophiliacs | | | 15 | 0 | 100.0% |
| RA (rheumatoid arthritis) | | | 10 | 0 | 100.0% |
| SLE (systemic lupus erythematosus) | | | 10 | 0 | 100.0% |
| ***Hepatic diseases:*** | | | | | |
| Alcohol induced hepatitis /cirrhosis | | | 13 | 0 | 100.0% |
| Drug induced hepatitis /cirrhosis | | | 10 | 0 | 100.0% |
| Fatty liver | | | 10 | 0 | 100.0% |
| Liver cancer | | | 10 | 0 | 100.0% |
| Non viral liver disease | | | 15 | 0 | 100.0% |
| **total** |  |  | **1468** | **0** | **100.0%** |

**Supplementary Table 2. Potential drug interferences in the ACOV2S assay.** Serum samples were spiked with drugs of interest and analyte recovery was calculated at three levels of analyte concentration: negligible (green), low (~0.5–4 U/mL, yellow) and high (~150–270 U/mL, blue).

|  |  | **Negligible analyte conc.** | **Low analyte conc.** | **High analyte conc** |
| --- | --- | --- | --- | --- |
|  |  | **Absolute deviation** | **Recovery (%)** | **Recovery (%)** |
| **Drug** | **Drug conc. (mg/L)** |  |  |  |
| **drugs with postulated potential to interrupt the RBD-ACE2 interface** | | | | |
| Risperidon | 0.03 | 0.000000 | 92.9 | 90.7 |
| Sitagliptin | 0.12 | 0.0132 | 92.7 | 90.8 |
| Baricitinib | 4.8 | 0.000000 | 99.1 | 91.1 |
| Silodosin | 9.6 | 0.000000 | 94.8 | 93.0 |
| Ebastin | 24 | 0.000000 | 94.7 | 94.1 |
| Indacaterolmaleat | 0.36 | 0.000000 | 94.9 | 99.0 |
| Regorafenib | 0.192 | 0.000000 | 89.8 | 90.8 |
|  | 0.064 | - | 97.5 | - |
| Omalizumab | 0.18 | 0.0012 | 99.4 | 98.0 |
| **special drugs used in COVID-19 treatment** | | | | |
| Zanamivir | 0.006 | - | 100.1 | 99.6 |
| Oseltamivir | 0.090 | - | 99.9 | 100.0 |
| Ceftriaxone | 2.40 | - | 98.0 | 96.7 |
| Levofloxacin | 0.300 | - | 101.2 | 100.3 |
| Meropenem | 3.60 | - | 101.1 | 100.0 |
| Ribavirin | 0.720 | - | 103.8 | 100.3 |
| Azithromycin | 0.300 | - | 104.2 | 99.4 |
| Arbidol | 0.120 | - | 100.6 | 97.3 |
| Lopinavir | 0.720 | - | 98.2 | 95.3 |
| α-interferon 2b | 3000 IE/mL | - | 100.0 | 99.1 |
| Peramivir | 0.360 | - | 98.4 | 98.8 |
| Tobramycin | 0.360 | - | 102.1 | 101.0 |
| Histamine Dihydrochl. | 0.0006 | - | 99.5 | 101.1 |
| Tocilizumab | 0.384 | - | 99.2 | 100.4 |
| α-interferon 2a | 43200 IE/mL | - | 102.2 | 100.9 |
| Hydroxychloroquinsulfat C1 | 0.240 | - | 96.0 | 99.5 |
| Remdesivir | 0.120 | - | 98.6 | 99.5 |
| Ritonavir | 0.480 | - | 74.5 | 68.4 |
|  | 0.360 | - | 80.1 | 76.0 |
|  | 0.240 | - | 89.5 | 85.1 |
|  | 0.160 | - | 95.5 | 92.4 |
|  | 0.120 | - | 95.9 | 96.0 |
| **Common** | | | | |
| Acetylcysteine | 150 | - | 100.8 | 100.0 |
| Acetylsalicylic acid | 30 | - | 102.6 | 102.7 |
| Ampicillin | 75 | - | 103.5 | 100.6 |
| Ascorbic acid | 52.5 | - | 102.8 | 101.3 |
| Cefoxitin | 750 | - | 101.9 | 101.3 |
| Doxycycline | 18 | - | 103.1 | 105.7 |
| Heparin | 3300 IU/L | - | 103.3 | 101.2 |
| Levodopa | 7.5 | - | 103.4 | 101.0 |
| Methyldopa | 22.5 | - | 103.2 | 100.3 |
| Metronidazole | 123 | - | 102.5 | 100.8 |
| Rifampicin | 48 | - | 103.3 | 100.0 |
| Acetaminophen | 156 | - | 100.4 | 98.6 |
| Cyclosporine | 1.8 | - | 100.9 | 91.8 |
| Ibuprofen | 219 | - | 100.3 | 92.9 |
| Theophylline | 60 | - | 100.3 | 90.7 |
| Phenylbutazone | 321 | - | 104.5 | 93.9 |
| Itraconazole | 7.5–30 | - | 87.8–93.7 | 86.9–95.7 |

**Supplementary Figure S1. Correlation of the ACOV2S assay with results from the cPass neutralization assay.** ACOV2S titers and percentage inhibition from the cPass assay from longitudinal samples from individual donors

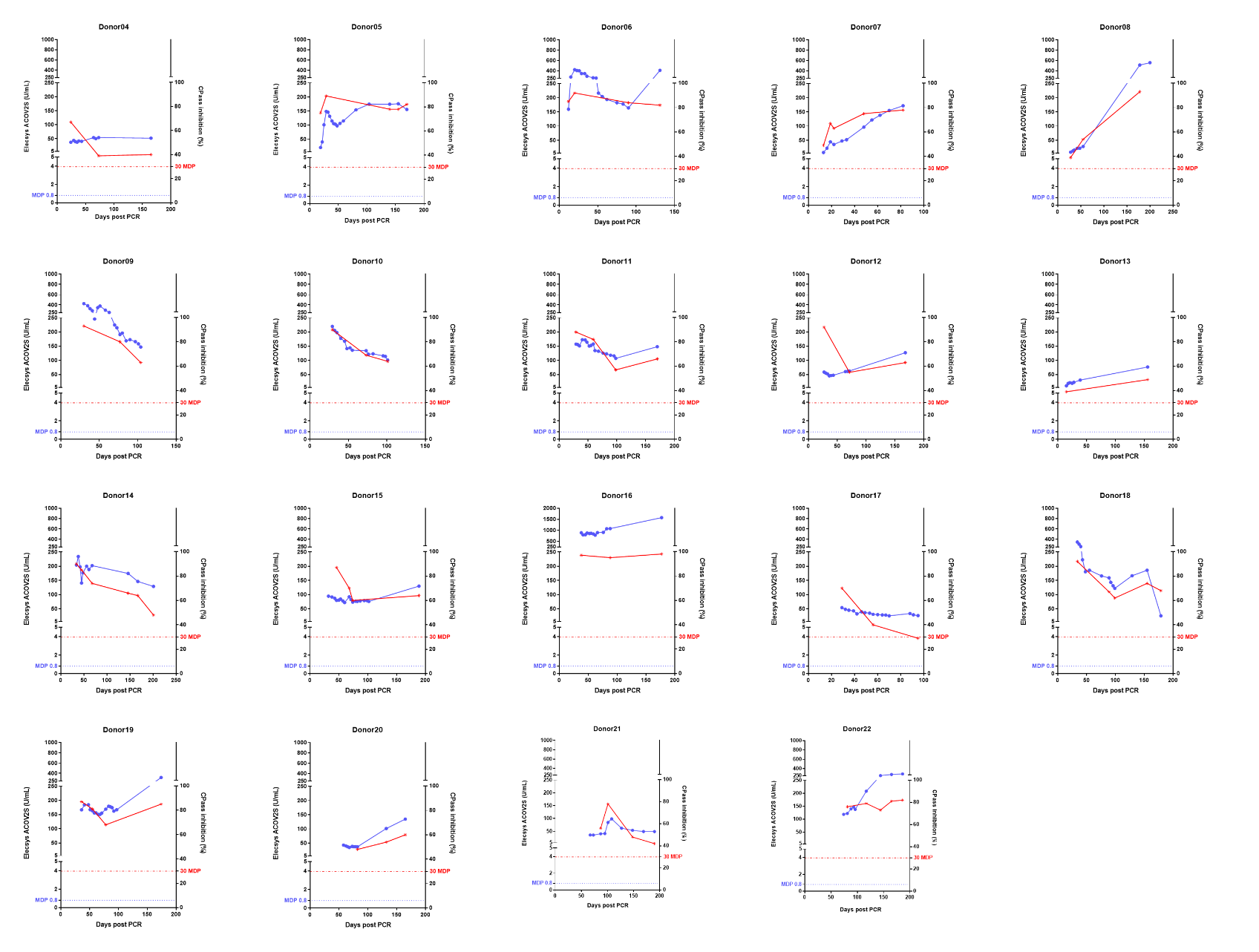

**Supplementary Figure S2. Correlation of the Genscript cPass and Elecsys ACE2-RBD neutralization assays.** (A) Method comparison with cutoffs marked (cPass 30%, ACE2-RBD, 25%), (B) residual plot, and (C) ROC curve. Red circles indicate samples from patients with severe COVID-19 (hospitalized) and blue triangles indicate samples from patients with mild COVID-19 (not hospitalized).

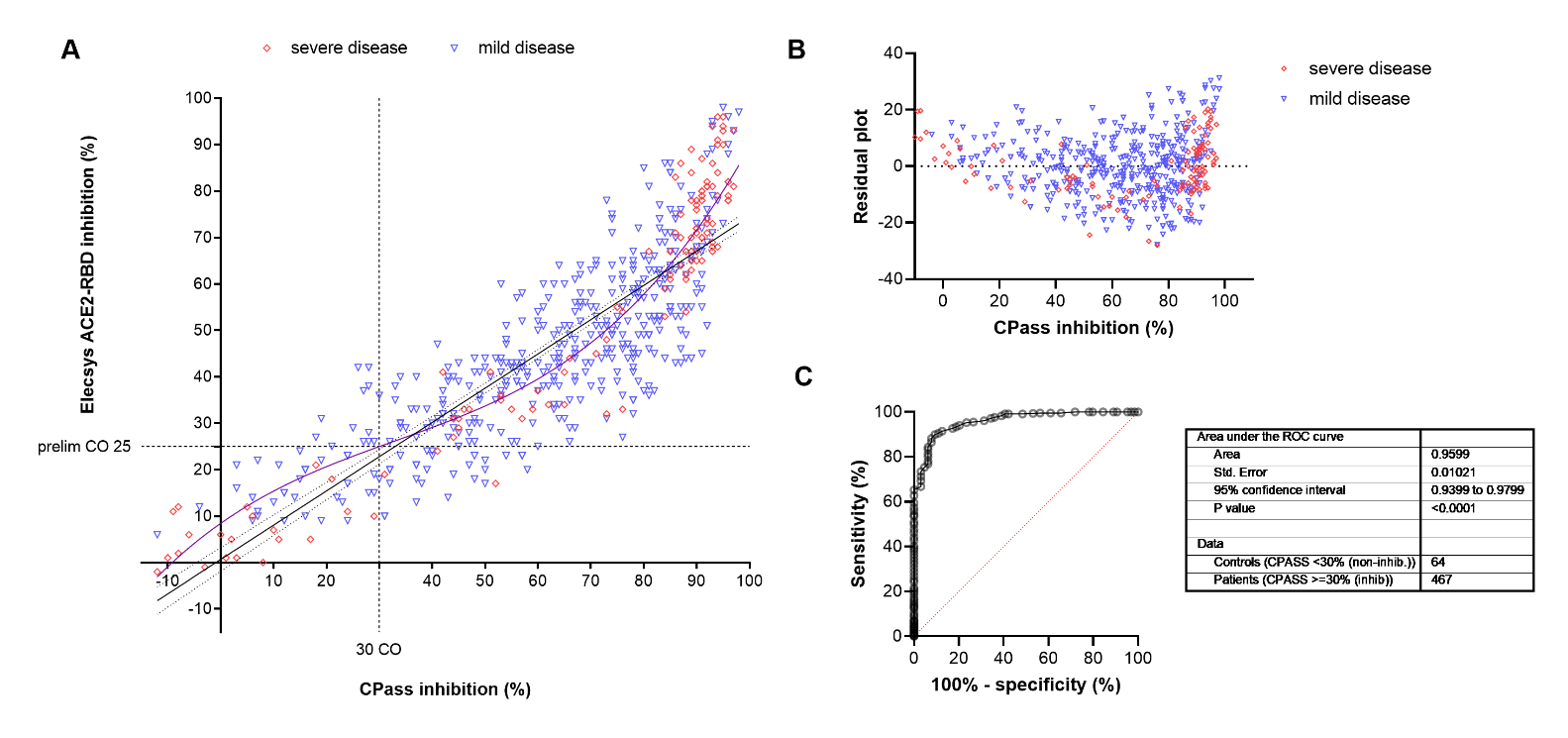

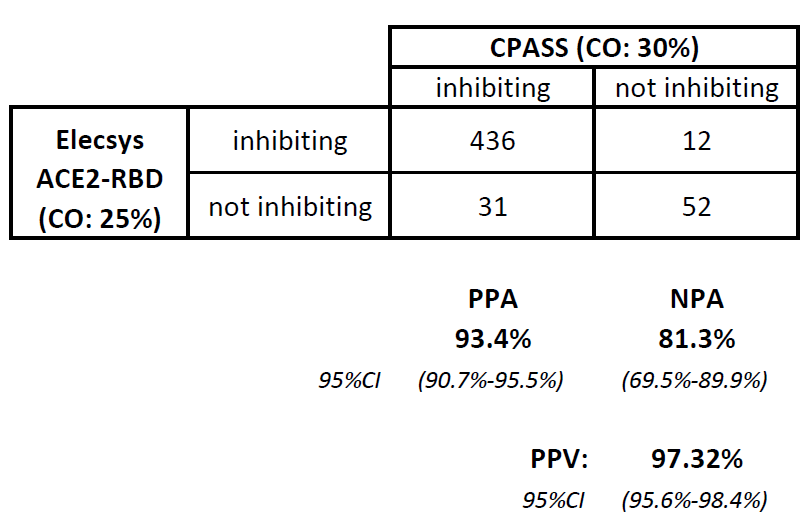
**D**.

**Supplementary Figure S3. Correlation of the Elecsys ACOV2S assay and Elecsys ACE2-RBS neutralization assay.** (A) Method comparison with cutoffs marked (ACOV2S 0.8 U/mL and 15 U/mL, ACE2-RBD, 25%), (B) residual plot, (C) ROC curve, and (D) qualitative agreement between the ACOV2S and ACE2-RBS assays. Red circles indicate samples from patients with severe COVID-19 (hospitalized) and blue triangles indicate samples from patients with mild COVID-19 (not hospitalized).

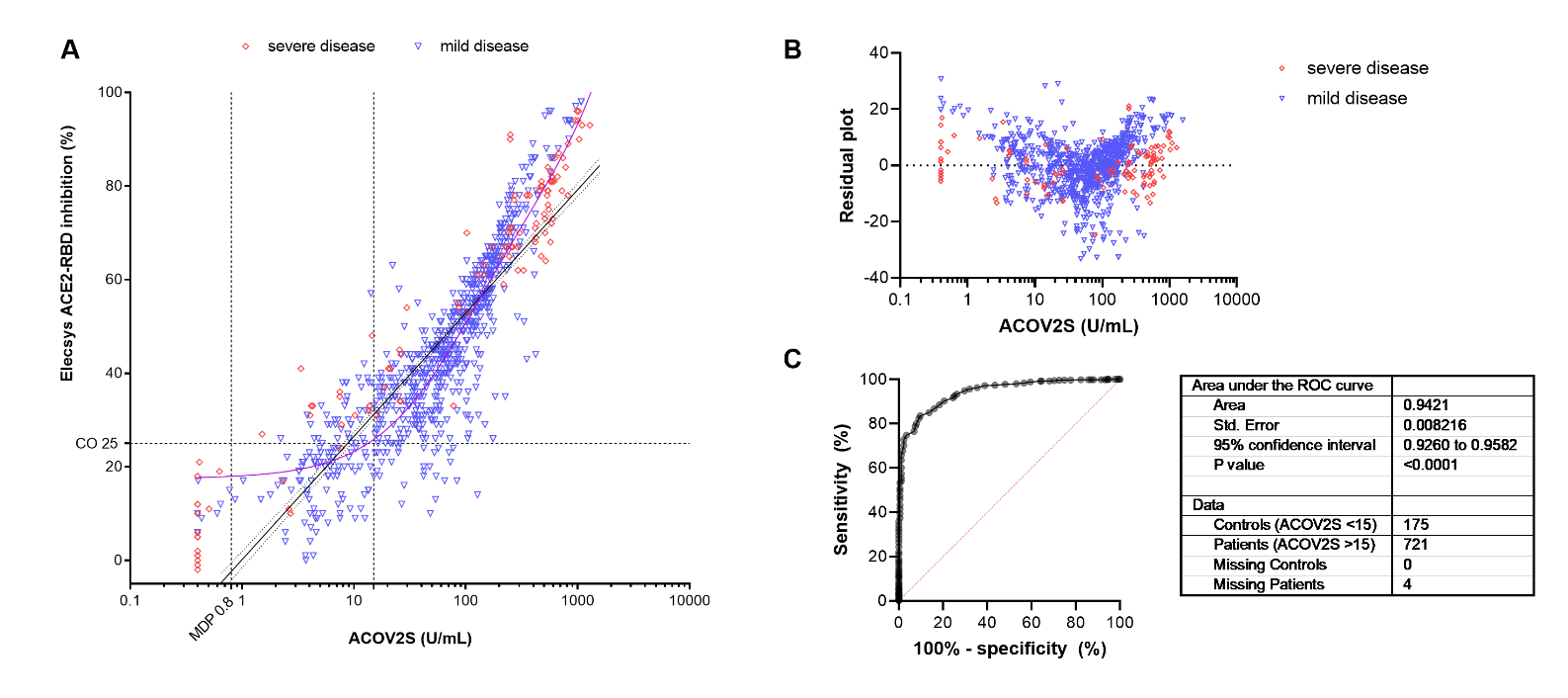

**D)**

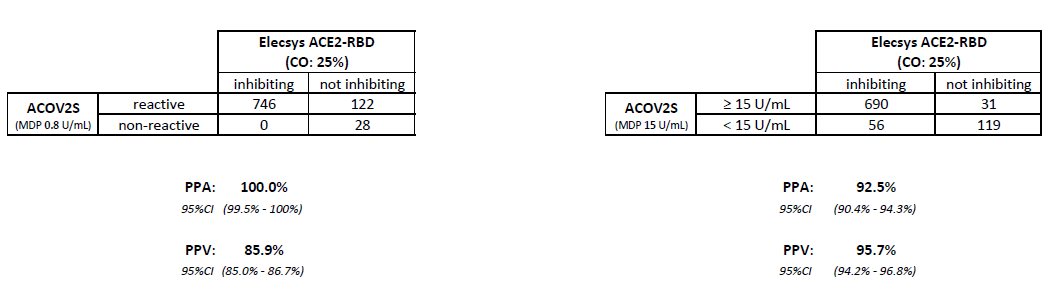
